# An application of machine learning to assist medication order review by pharmacists in a health care center

**DOI:** 10.1101/19013029

**Authors:** Maxime Thibault, Denis Lebel

## Abstract

The objective of this study was to determine if it is feasible to use machine learning to evaluate how a medication order is contextually appropriate for a patient, in order to assist order review by pharmacists. A neural network was constructed using as input the sequence of word2vec embeddings of the 30 previous orders, as well as the currently active medications, pharmacological classes and ordering department, to predict the next order. The model was trained with data from 2013 to 2017, optimized using 5-fold cross-validation, and tested on orders from 2018. A survey was developed to obtain pharmacist ratings on a sample of 20 orders, which were compared with predictions. The training set included 1 022 272 orders. The test set included 95 310 orders. Baseline training set top 1, top 10 and top 30 accuracy using a dummy classifier were respectively 4.5%, 23.6% and 44.1%. Final test set accuracies were, respectively, 44.4%, 69.9% and 80.4%. Populations in which the model performed the best were obstetrics and gynecology patients and newborn babies (either in or out of neonatal intensive care). Pharmacists agreed poorly on their ratings of sampled orders with a Fleiss kappa of 0.283. The breakdown of metrics by population showed better performance in patients following less variable order patterns, indicating potential usefulness in triaging routine orders to less extensive pharmacist review. We conclude that machine learning has potential for helping pharmacists review medication orders. Future studies should aim at evaluating the clinical benefits of using such a model in practice.

## 1 Introduction

In North American health care centers, medication orders are reviewed by pharmacists to ensure that there are no drug related problems or errors which could cause suboptimal care or even harm.[1–3] Within health-system pharmacy practice, this activity falls under the scope of pharmacy services, which may be defined as all activities that involve processing medication orders, preparing and dispensing medication. Another aspect of pharmacist activity in health systems is pharmaceutical care, or clinical pharmacy, which encompasses the provision of direct care to patients in collaboration with other health care professionals and providers. The profession of health-system pharmacy has been undergoing a shift from a services focused profession towards a primary focus on clinical pharmacy.[4]

### 1.1 Medication order review

The activity of drug order review has generally been regarded as a crucial safeguard to ensure the safe use of medication. While it would not be professionally acceptable to prepare and dispense drugs without verifying that the orders for these drugs are appropriate, modern evidence supporting the safety benefits of pharmacist order review in hospitals is surprisingly scarce.[1–3]

In most centers, medication order review is centralized. Experience shows that decentralizing order verification to clinical pharmacists is detrimental to their ability to perform timely clinical interventions. However, resources allocated to these tasks are interconnected as clinical pharmacy activities cannot be expanded if pharmaceutical services are not adequately covered.

With the generalized implementation of electronic health records and computerized provider order entry, pharmacists have gained immediate access to all orders and pertinent patient data. This led to an impetus for pharmacists to prospectively review all medication orders. This is enabled by technology such as automated dispensing cabinets, which have the capability to allow access to a drug only when a pharmacist has verified an order. In this context, drug order verification is now sometimes called nearly universal prospective order review (NUPOR).[5]

While NUPOR may be regarded as a necessity for the safe use of drugs in hospitals, critics have pointed out that performing order review prospectively to ensure that drugs are available in a timely manner puts pressure on pharmacists and pharmacy departments to assign resources to this activity, which may negatively impact pharmacist availability to perform pharmaceutical care.[5–7] Additionally, pharmacists have observed that in the context of computerized provider order entry with clinical decision support, a large majority of orders contain no errors.[8] More recent data reminds that pharmacists are not infallible. Indeed, as the number of orders verified by a pharmacist during a shift increase, so does the risk of committing an error.[9] The areas where orders are verified are busy work zones, and pharmacists performing review must often interrupt their work to perform other tasks like answering phone calls. A simulation study by our group showed that a pharmacist being interrupted while verifying an order has a decreased chance of detecting an error within that order.[10]

We believe that it would be desirable to move away from the concept of NUPOR and focus pharmacist attention on orders which are more likely to contain errors, that is orders that are unusual in some way, instead of prospectively verifying every single order. However, a challenge lies in the identification of these orders, which should ideally be done without human intervention. Recent advances in machine learning could help with this challenge.

### 1.2 Using machine learning to analyze prescriptions

The analysis of drug orders using machine learning would require two steps of verification. First, the prescribed dose, route of administration and frequency of administration would need to be analyzed in relation with the drug. Second, the medication itself would need to be analyzed in context of the patient’s current situation.

This first element of verification was studied by Woods et al. [11], who used the statistical frequency of a combined representation of dose, route and frequency, in relation to a pharmaceutical product, to determine if a specific prescription is atypical for a selection of five drugs. Flynn et al. [12] expanded this approach in 2018 to 431 drugs prescribed to patients aged 75 and over. This study showed that, defining as “rare” a dose, route and frequency combination occurring in <= 10% of cases on a training set, only 27.3% of orders would be considered rare in their testing set.

In order to perform the second verification, one would need to determine if a prescribed medication is typical or not, given a patient’s context. A machine learning model trained on past medication orders could learn order patterns and could be able to warn a pharmacist if a particular order is unusual in a given context. This could involve predicting contextually likely medication orders and determining if an actual order fits within those predictions.

In 2008, Helgason [13] used sequential pattern mining to predict the next prescription, defined as the generic drug name, and showed 70.2% top 20 accuracy among 978 possibilities. More recently, Wright et al. [14] also used sequential pattern mining and succeeded at predicting the next drug class and the next generic drug name that would be prescribed to outpatients with diabetes with respectively 90% and 64.1% accuracy. Chen et al. [15] evaluated how order sets, as compared to latent Dirichlet allocation probabilistic topic modeling, predicted clinical orders within 24 hours. Order sets proved less effective at this prediction than the probabilistic topic model with respective area under the receiver operating characteristic curves (AUROCs) of 0.81 and 0.90. These data show that order patterns can be learned by algorithms and used for predictions. However, no study has applied these predictions to the context of medication order review by pharmacists.

### 1.3 Study objectives

The primary objective of this study was to determine if it is feasible to use machine learning to evaluate how well a medication order fits a patient’s context, in a setting of pharmacist order verification. Secondary objectives were to evaluate the performance of the resulting model across patient populations, and to determine if the predictions of the model correlated with how atypical pharmacists rated a sample of orders as a measure of clinical performance.

## 2 Methods

### 2.1 Setting

This study was performed with data extracted from the pharmacy database of CHU Sainte-Justine, a 500-bed mother and child university hospital center located in Montréal, Canada. Access to the data was authorized in conformity to local requirements.

### 2.2 Dataset

Extraction of the data was performed in August 2018. A training set consisting of all orders entered between 2013 and 2017 inclusively was constituted, as well as a test set of all orders between January 2018 and July 2018 inclusively. Data was cleaned up to remove canceled orders, obvious entry errors (e.g. orders entered in 2013 with a start date in 2031) as well as fictional patients which exist for testing or other purposes.

The extracted data included a de-identified patient encounter ID, the date of admission, the drug identifier and name, the AHFS pharmacological class of the drug, the order start and end times precise to the minute, as well as the department on which the patient was when the order was entered.

### 2.3 Preprocessing

The data was preprocessed so that each individual order was considered as a label. The features associated with this label were:

- The sequence of up to 30 drug orders preceding the label.
- The active drugs at the time the label was entered.
- The active pharmacological classes at the time the label was entered.
- The department on which the patient was when the label was entered.

Because the AHFS class is a four-level hierarchical class, the four levels were decomposed and included into the features to provide as much information as possible to the model. For example, a drug having an AHFS class of 08:12.06.04 would be decomposed into 08, 08:12, 08:12.06 and 08:12.06.04 and these four elements would be included into the active pharmacological classes feature.

### 2.4 Representation of medication order sequences as word2vec embeddings

The sequence of 30 drug orders preceding the label can be thought of as a “sentence” carrying meaning. For example, a sequence going from oral medications to intravenous medications to intravenous infusions could indicate a patient whose clinical condition is deteriorating. For this reason, we turned to natural language processing techniques, specifically word2vec, to represent this feature.

Word2vec is a natural language processing technique that produces word embeddings by using a shallow neural network to predict a word from the words that come immediately before it and after it in a sentence (the continuous bag of words (CBOW) approach) or, conversely, to predict the words before and after from the word itself (the skip-gram approach). This technique has been applied to a variety of subjects in biomedical sciences with interesting results.[16–22] A hallmark feature of word2vec is that the resulting embeddings encode semantic relationships. A classic example would be that, in embeddings trained on an English language corpus, the vectors going from countries to their capital are similar.[23]

In the context of medical data, a modified word2vec approach called cui2vec has been applied to insurance claims, clinical notes and biomedical journal articles to extract and encode clinical concepts.[24] The authors evaluated this model by determining how well “may treat” and “may prevent” relationships between drugs and conditions were represented, and if drugs belonged to the same semantic type. However, the prediction of medication order sequences using this technique was not evaluated, as the claims data used in this experiment did not contain drugs.

Another modified approach called med2vec has been used to model diagnostic, procedural and medication codes from patient visits.[25] The authors used these med2vec embeddings to predict codes for future visits, but not as a sequence within a single hospitalization. Additionally, they noted that the technique did not perform as well for medications as for other codes.

Given that these two modified techniques were not readily applicable to our clinical problem without further tuning and evaluation, we opted to train “regular” word2vec embeddings on the raw sequences of orders for every encounter.

We used two techniques to evaluate the embeddings. We first composed a list of 153 analogies between different medications, for example “acetaminophen 80 mg/mL oral solution is to acetaminophen 325 mg tablet as to furosemide 10 mg/mL oral solution is to× “. We trained a word2vec model using random search and then grid search with 3-fold cross validation to find the best hyperparameters to maximize the accuracy on this list of analogies. These best hyperparameters were then used in the word2vec training pipeline that was used during cross-validation of the entire neural network.

Secondly, we projected the embeddings into three dimensions using the UMAP technique [26] and performed agglomerative clustering on the resulting projection. Grid search with 3-fold cross-validation was used to determine the number of clusters that maximized the silhouette score of the clusters, and the resulting clusters were labeled by a pharmacist familiar with drugs used at this institution (MT) to evaluate clinical concept modeling.

### 2.5 Representation of active drugs as a multi-hot vector

Active drugs and classes and the time of entry of the label are lists of variable length among a fixed set of possibilities. We represented these lists as a bag of words, wherein the lists become unordered collections of “words” representing the drugs and the four levels of classes. The ordering department was also included into this bag of words. Binary count vectorization was performed on these bags of words to transform them into multi-hot vectors of fixed length.

### 2.6 Neural network

The sequence of word2vec embeddings as well as the multi-hot vector were used as two inputs to a neural network classifier. The sequences were left-padded with zeros when less than 30 orders were present before the label.

For the neural network, we performed all experiments with 5-fold cross-validation using a time-series split. This splitting strategy was used to allocate encounters to either a training or validation split at each fold, such that each training split would only include encounters that began before the encounters allocated to the corresponding validation split. This allowed preservation of the temporal relationship between the training and validation splits, specifically to avoid leaking information about future clinical practices into the training set.

To further avoid information leakage, at every fold during cross-validation, the word2vec embeddings and the multi-hot vector pipeline were retrained using only the training data for that fold. The Adam optimizer was used, and categorical cross-entropy was used as the loss function. During training, the learning rate was reduced by a factor of 10 when validation loss decreased less than 0.0005 after 3 epochs and early stopping occurred then validation loss decreased less than 0.0001 for 5 epochs despite learning rate reduction.

Experiments were performed to find the best network configuration and training hyperparameters. After the final model configuration was determined, we created two modified models by ablating one of the two inputs, to determine if both inputs were useful. 5-fold cross-validation was used to compare the performance of single-input models against the two-input model. After all experiments were done, a final model was fitted using the entire training set and tested against the 2018 set.

For the test set, we computed the precision and recall of each drug prescribed more than once a week, and we also computed the weighted average precision, recall and AUROC over the entire test set as measures of classification performance. Finally, we computed the accuracy measures as well as weighted average precision and recall by department as a proxy category for patient population.

### 2.7 Correlation with pharmacist rating of orders

The final model was deployed as a prototype on the institutional intranet, showing pharmacists, in a simplified fashion using color cues, where each medication prescribed for a patient ranked in the model predictions at the time of prescription. Orders representing 20 clinical situations (e.g. routine order, possible but unusual combination) were sampled from this prototype. Four of these orders were modified to simulate an order entry error. We constructed an online survey for which we solicited participation from all pharmacists working at our center and not involved in this project. We asked participants to rate on a four-point scale, given the same information as the model, to which degree they found that each order was atypical. Correlation between pharmacist ratings was verified using the Fleiss kappa.

The correlation between binned prediction ranks and pharmacist ratings was evaluated using the Cohen kappa, and we computed the precision, recall and accuracy of the binned model predictions as compared to pharmacist ratings.

### 2.8 Software and code

The following Python packages were used primarily to perform this study: Gensim [27], Scikit-Learn [28], Tensorflow [29], UMAP [26]. The full code used for this study can be found on GitHub. We provide the preprocessor we used which can be adapted to medication data from other sources. We also provide a working example adapted to the MIMIC-III dataset.[30] The results obtained with MIMIC-III should be interpreted with caution. MIMIC-III includes only data from ICU stays while our data included ICU stays but is not limited to them. The MIMIC-III medication orders are precise to the day, while our data was precise to the minute, which impairs the representation of drug orders as a sequence. Finally, the MIMIC-III dataset does not include pharmacological classes. This example performs poorly and should only be used as a concept demonstration and not as a valid experiment.

## 3 Results

### 3.1 Dataset

The training set included 1 022 272 orders for 3145 drugs from 96 590 encounters, with (mean ± sd) 11.3± 22.7 orders per encounter. The testing set included 95 310 orders for 1843 drugs from 95 310 encounters, with (mean ± sd) 10.2 ±15.3 orders per encounter.

### 3.2 Word2vec embeddings

The best training hyperparameters for word2vec embeddings were an embedding dimension of 128 with the CBOW method, negative sampling, a window of 5, and a learning rate of 0.013 trained for 32 epochs. This yielded an analogy accuracy of 77.1% (118/153). Grouping the 3D UMAP-projected embeddings into 37 clusters yielded a (mean± sd) silhouette score of 0.470 ± 0.021. The resulting embeddings and color-coded clusters are shown in Figure 1. Four clusters could not be labeled to a specific concept. Other clusters corresponded to clinical concepts of various nature:

- Departments (e.g. neonatology, intensive care)
- Medical specialties (e.g. psychiatry, dermatology)
- Types of pharmaceutical products (e.g. vaccines, wound care)
- Clinical conditions (e.g. tuberculosis, *hyperemesis gravidarum*)

**Figure 1:**
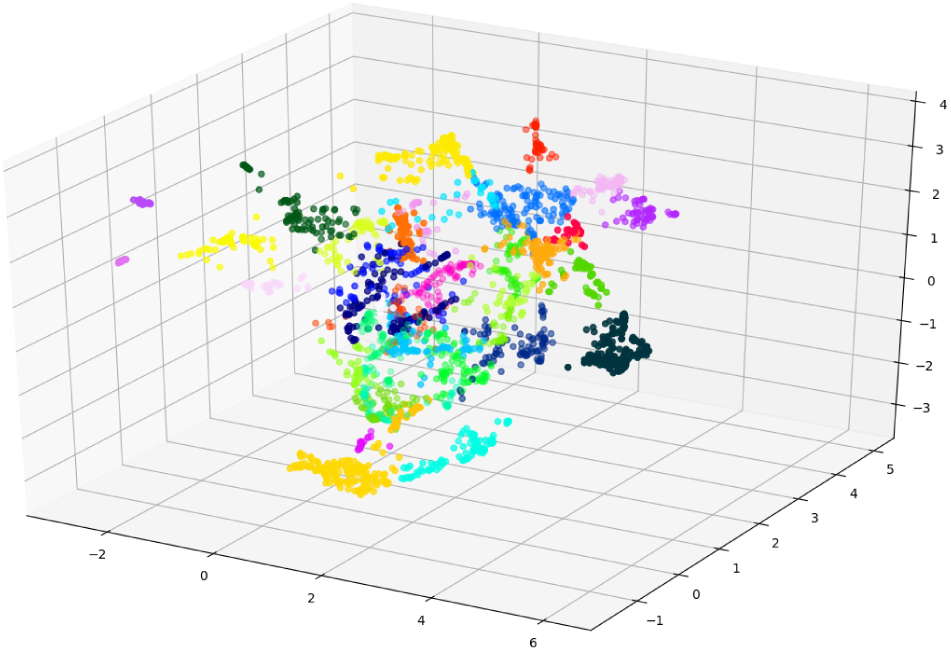
UMAP 3D projection of clustered word2vec embeddings. Colors indicate clusters.

### 3.3 Neural network

The final structure of the neural network is shown in Figure 2. The LSTM layers and the dense layers used for the sequence had 128 nodes. The fully-connected layer used for the multi-hot vector had 128 nodes. The concatenation layer as well as the fully-connected layer up until output had 256 nodes. The best hyperparameters were a dropout rate of 0.2 and a batch size of 256. For the final model, training was performed for 7 epochs. The prediction accuracy of the model can be found in Table 1. Combining the two inputs proved superior to using either input alone, as shown in Figure 3.

**Table 1:** Prediction accuracy

|  | Top 1 (%) | Top 10 (%) | Top 30 (%) |
| --- | --- | --- | --- |
| Baseline on training set<sup>1</sup> | 4.5 | 13.6 | 44.1 |
| Cross-validation accuracy (mean ± sd) | 44.0 ± 0.4 | 68.2 ± 1.3 | 78.8 ± 1.3 |
| Test set accuracy | 44.4 | 69.9 | 80.4 |

<sup>1</sup>Frequency-based classifier, always predicts the most common top 1, top 10 and top 30 drugs

**Figure 2:**
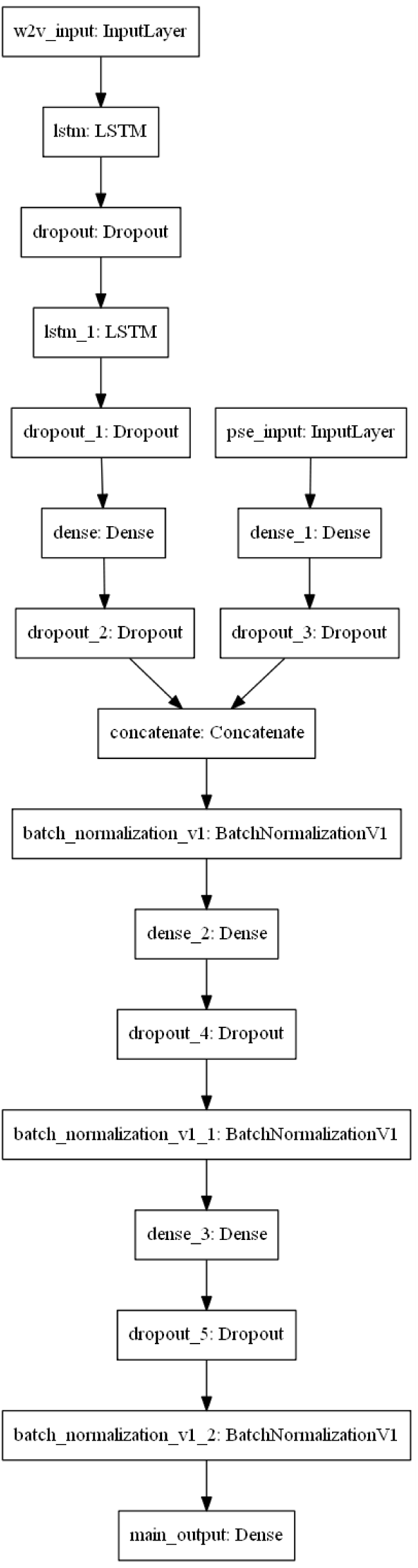
Final neural network structure

**Figure 3:**
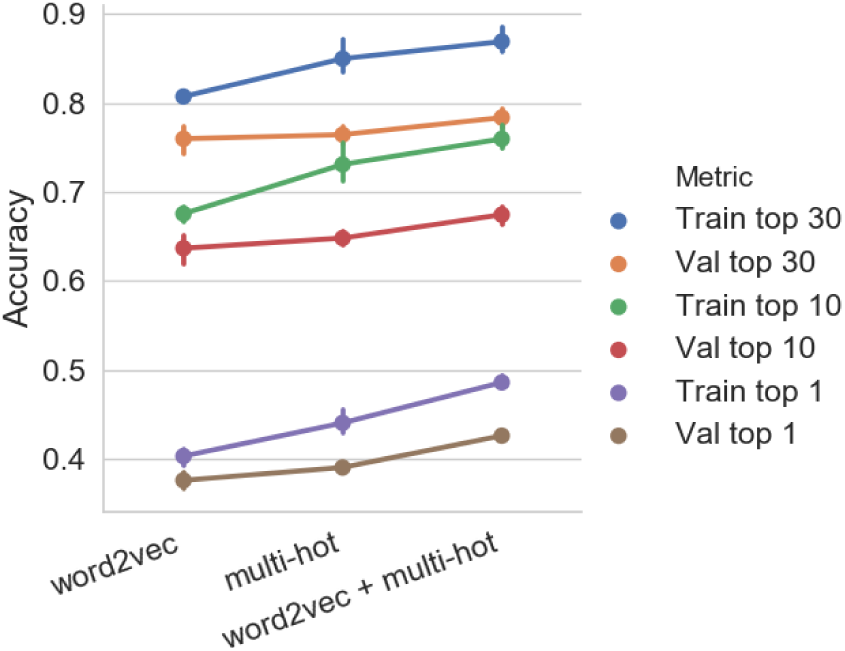
Validation performance of single and dual input models

On the 2018 test set, 264 (0.3%) orders were discarded because of previously unseen labels. The weighted average precision, recall and AUROC were respectively 0.416, 0.444 and 0.959. 1176 different drugs were predicted by the model. Individual precision and recall values for drugs prescribed more than once a week can be seen in Figure 4. Performance by department is shown in Table 2.

**Table 2:** Prediction performance on test set by department

|  | Orders (n) | Unique drugs prescribed (n) | Top 1 accuracy (%) | Top 10 accuracy (%) | Top 30 accuracy (%) | Precision<sup>1</sup> | Recall<sup>1</sup> |
| --- | --- | --- | --- | --- | --- | --- | --- |
| Adolescents | 229 | 78 | 14.4 | 41.5 | 55.9 | 0.091 | 0.144 |
| Ambulatory clinics | 2 | 2 | 100 | 100 | 100 | 1 | 1 |
| Emergency | 148 | 102 | 11.5 | 39.9 | 62.2 | 0.117 | 0.115 |
| General pediatrics | 15 581 | 992 | 27.8 | 58.5 | 72.5 | 0.238 | 0.278 |
| Long term care | 1154 | 311 | 9.6 | 30.9 | 48.1 | 0.079 | 0.096 |
| Neonatal intensive care | 4536 | 262 | 46.9 | 77.3 | 89.1 | 0.417 | 0.469 |
| Newborns | 332 | 47 | 55.7 | 84.9 | 91.3 | 0.521 | 0.557 |
| Obstetrics / gynecology | 32 637 | 563 | 71.1 | 87.3 | 92.0 | 0.707 | 0.711 |
| Oncology | 12 665 | 740 | 28.0 | 56.9 | 71.1 | 0.245 | 0.280 |
| Outpatients | 2 | 2 | 100 | 100 | 100 | 1 | 1 |
| Pediatric intensive care | 2724 | 480 | 18.3 | 43.5 | 59.9 | 0.150 | 0.183 |
| Psychiatry | 535 | 152 | 18.3 | 43.7 | 64.1 | 0.102 | 0.183 |
| Specialized pediatrics | 11 173 | 885 | 29.0 | 59.6 | 73.0 | 0.248 | 0.290 |
| Surgery | 13 592 | 756 | 36.2 | 69.2 | 80.9 | 0.320 | 0.362 |

<sup>1</sup>Weighted average

**Figure 4:**
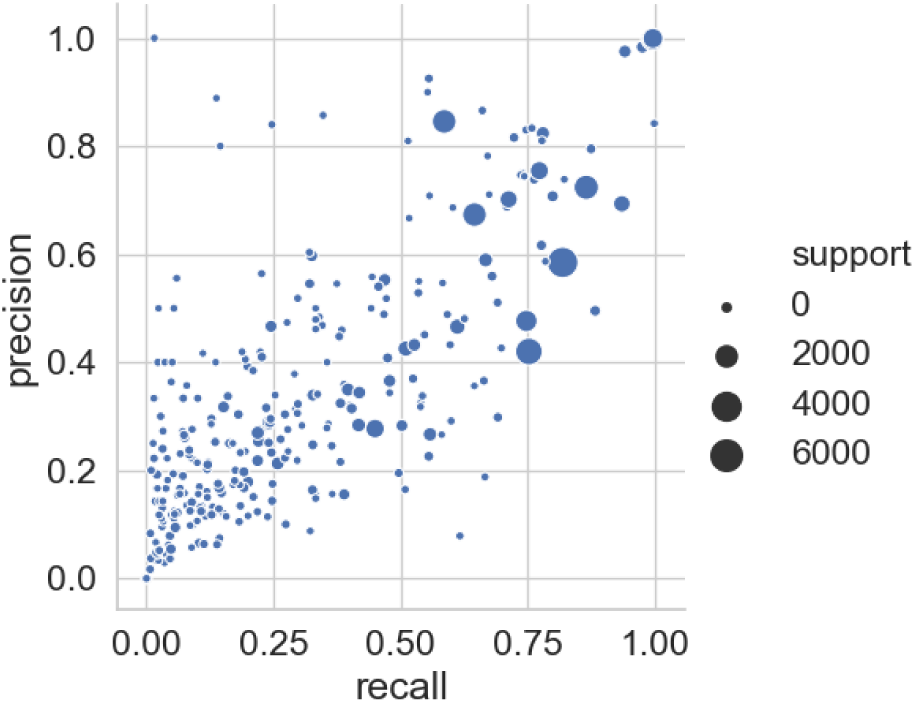
Precision and recall on test set for drugs prescribed more than once a week

### 3.4 Correlation with pharmacist rating of orders

18/35 (51.4%) pharmacists answered the survey. The correlation between pharmacist ratings was poor with a Fleiss kappa of 0.283. Correlation between the pharmacist ratings and the binned model predictions is shown in Table 3.

**Table 3:** Correlation of predictions with pharmacist ratings

|  | Accuracy | Cohen kappa | Precision<sup>1</sup> | Recall<sup>1</sup> |
| --- | --- | --- | --- | --- |
| Overall<sup>2</sup> | 0.550 | 0.338 | 0.617 | 0.550 |
| Real orders<sup>3</sup> | 0.594 | 0.334 | 0.679 | 0.594 |

<sup>1</sup>Weighted average<sup>2</sup>20 orders<sup>3</sup>Excluding simulated errors, 16 orders

## 4 Discussion

Our results show that is it feasible to predict the next medication order during hospitalization using a neural network classifier.

We observed that the model performed much better on certain populations of patients (obstetrics and gynecology, newborns, neonatal intensive care patients) than on others. The features contained in our dataset did not allow for systematic exploration of the reasons behind this variation. However, our experience as pharmacists reviewing orders for these patients tells us that they generally have a relatively similar clinical background. As a result, these patients will generally follow similar ordering patterns unless some unusual element (e.g. rare comorbidity, complication) is present in their context of care. Patients in other categories have more heterogenous conditions and can follow more varied ordering paths.

While the performance on certain populations of patients may seem discouraging, this should be interpreted in the context of the aim of such a model, which would be to triage routine orders to a less exhaustive review, with the ultimate goal of requiring prospective, extensive pharmacist review only for unusual orders. The fact that the model seems better at predicting orders for the populations which we find more likely to follow a pattern of routine orders is in line with this expected behaviour, and in our opinion, is an interesting finding.

Critics of NUPOR have specifically called out routine post-childbirth orders, which are common in our obstetrics patients, as a priority target for triage to a less extensive order review process.[6] This appears to us as an interesting avenue to free up pharmacist time to provide more value-added pharmaceutical care to this population. Similarly, medical orders for newborn babies who are not admitted to the neonatal intensive care unit (NICU) are generally very straightforward. Spending less time reviewing orders for these patients could offer an opportunity to provide better pharmaceutical care to sicker babies admitted to the NICU.

Comparison with pharmacist rating of orders proved difficult. Pharmacists were not in agreement about how unusual the sampled orders were, which is a known issue with data labeling by experts.[31] A variety of factors could explain this issue. All pharmacists practicing at our center were invited to rate the same orders, and as such, each individual pharmacist needed to rate several orders outside of his or her field of clinical expertise. Additionally, pharmacists were provided with the same information as the model, without a specific clinical scenario or patient information, which is not a realistic simulation of practice. The participants may have rated how clinically appropriate they found each order rather than how typical or atypical it was. Indeed, some pharmacists who completed the survey told us that they did not realize that the goal was not to find clinically inappropriate orders. This method for evaluating the clinical performance of the model may not be optimal. Further studies evaluating the practical usefulness of such a model should use a different method.

There is an inherent randomness in the ordering sequence of medications. When a provider places multiple orders in a short time period (i.e. entering a group of orders one at a time), they may not enter them in a predictable sequence. It would be unreasonable to expect the model to predict the exact next medication with high accuracy in this context, which is why top 10 and top 30 accuracies were used as more realistic measures, as was done in the med2vec report.[25] In practical use, several predictions (the exact number would likely vary by population) would be obtained from the model at the time an order is received and these predictions would then be compared to the order to determine if an order is atypical.

The only other study which reported similar metrics over a wide spectrum of medications prescribed to inpatients is the thesis by Helgason [13]. The best model in this study showed 70.2% top 20 accuracy. The number of orders and encounters included in that dataset was comparable to what was used in our study. However, the dataset in the Helagson study was aggregated from multiple hospitals, provided prediction only at the generic drug level, and included mostly older patients (mean age 58.7 years), while the data from our study is from a single mother-and-child university hospital and provided predictions at the level of the exact medication entry. The results in the Helagson study were not broken down by patient population. Still, the fact that comparable global prediction performance was observed indicates that this approach may be feasible over a variety of patients.

As stated before, the limits of this study include the fact that it is limited to a single center. We are currently setting up collaborations to test this technique in other hospitals. Although one study showed comparable overall metrics, future studies in other centers should also report performance in specific populations to allow for more granular comparison. Our model provides a prospective evaluation for a single order, at the time this order is reviewed by a pharmacist. We believe it would be useful to allow for a retrospective analysis of all currently active orders that would take into account all information available at the time of analysis, and not only information temporally preceding each order. An model building on the results presented in this paper is currently being developed for this task. Finally, we did not evaluate the clinical impact of using the model prototype in practice, which we plan to do in the future.

A limitation of our approach is that to provide meaningful feedback to a pharmacist verifying orders, the label needs to be precise at the level of the exact drug entry. This is required because a single drug may exist as several software entries representing different pharmaceutical products which may be appropriate only in certain contexts. For example, acetaminophen oral solution would be appropriate for neonates but not acetaminophen tablets. Similarly, a single drug product may exist as general or context specific entries. In our system build, specific aminoglycoside entries exist for neonatal patients. These entries include different administration instructions and schedules for nursing staff. A prediction model aiming to detect contextually inappropriate entries needs to differentiate between these, which is impossible if the label is encoded at the generic drug level. Because drug product databases are seldom shared or standardized between institutions, this requirement makes aggregation of sufficiently detailed data between centers difficult. In this context, the best approach for practical application of such a model would probably be to train models on data from individual institutions and to aim for a generalizable training method, instead of a single generalizable model.

## 5 Conclusion

This study showed that it is feasible to train a neural network that learns medication order patterns, with the aim of comparing actual orders to predictions, to assist pharmacists in determining if an order is atypical in a patient’s context. Prediction performance varied across patients populations, with obstetrics and gynecology patients and newborn babies being the categories where the model showed the best results. The rating of sampled orders by pharmacists to compare pharmacist opinion with model predictions was shown to be unreliable, with poor agreement between pharmacists. Future research should try other methods to provide more robust metrics of clinical performance.

## Data Availability

The dataset used for this study may be accessed under a data sharing agreement. Please contact the authors for more information.

## References

[1] Sara Ibáñez-Garcia, Carmen Guadalupe Rodriguez-Gonzalez, Maria Luisa Martin-Barbero, Maria Sanjurjo-Saez, Ana Herranz-Alonso, and iPharma. Adding value through pharmacy validation: A safety and cost perspective. Journal of Evaluation in Clinical Practice, 22(2): 253–260, April 2016. doi: 10.1111/jep.12466.

[2] B. Charpiat, S. Goutelle, M. Schoeffler, F. Aubrun, J.-P. Viale, C. Ducerf, G. Leboucher, and B. Allenet. Prescriptions analysis by clinical pharmacists in the post-operative period: A 4-year prospective study. Acta Anaesthesiologica Scandinavica, 56(8):1047–1051, September 2012. doi: 10.1111/j.1399-6576.2011.02644.x.

[3] Adnan Beso, Bryony Dean Franklin, and Nick Barber. The frequency and potential causes of dispensing errors in a hospital pharmacy. Pharmacy world & science: PWS, 27(3):182–190, June 2005. doi: 10.1007/s11096-004-2270-8.

[4] Craig A. Pedersen, Philip J. Schneider, Michael C. Ganio, and Douglas J. Scheckelhoff. ASHP national survey of pharmacy practice in hospital settings: Monitoring and patient education—2018. American Journal of Health-System Pharmacy, 76(14):1038–1058, July 2019. doi: 10.1093/ajhp/zxz099.

[5] Allen J. Flynn. Opportunity cost of pharmacists’ nearly universal prospective order review. American Journal of Health-System Pharmacy, 66(7):668–670, April 2009. doi: 10.2146/ajhp070671.

[6] John Poikonen. An informatics perspective on nearly universal prospective order review. American Journal of Health-System Pharmacy, 66(8):704–705, April 2009. doi: 10.2146/ajhp080410.

[7] Dennis A. Tribble. Automating order review is delegation, not abdication. American Journal of Health-System Pharmacy, 66(12):1078–1079, June 2009. doi: 10.2146/ajhp090095.

[8] Charles D. Mahoney, Christine M. Berard-Collins, Reid Coleman, Joseph F. Amaral, and Carole M. Cotter. Effects of an integrated clinical information system on medication safety in a multi-hospital setting. American Journal of Health-System Pharmacy, 64(18):1969–1977, September 2007. doi: 10.2146/ajhp060617.

[9] Christy Gorbach, Linda Blanton, Beverly A. Lukawski, Alex C. Varkey, E. Paige Pitman, and Kevin W. Garey. Frequency of and risk factors for medication errors by pharmacists during order verification in a tertiary care medical center. American Journal of Health-System Pharmacy, 72(17): 1471–1474, September 2015. doi: 10.2146/ajhp140673.

[10] Maxime Thibault, Céline Porteils, Stéphanie Goulois, Arielle Lévy, Denis Lebel, and Jean-François Bussières. The Impact of Phone Interruptions on the Quality of Simulated Medication Order Validation Using Eye Tracking: A Pilot Study. Simulation in Healthcare, 14(2):90–95, April 2019. doi: 10.1097/SIH.0000000000000350.

[11] Allie D Woods, David P Mulherin, Allen J Flynn, James G Stevenson, Christopher R Zimmerman, and Bruce W Chaffee. Clinical decision support for atypical orders: Detection and warning of atypical medication orders submitted to a computerized provider order entry system. Journal of the American Medical Informatics Association, 21(3): 569–573, May 2014. doi: 10.1136/amiajnl-2013-002008.

[12] Allen J. Flynn, Julia Adler Milstein, Peter Boisvert, Nate Gittlen, Carl Lagoze, and George Meng. ScriptNumerate: A Data-to-Advice Pipeline using Compound Digital Objects to Increase the Interoperability of Computable Biomedical Knowledge. AMIA Annual Symposium Proceedings, 2018:440–449, December 2018.

[13] Ívar Sigurjón Helgason. Predicting Prescription Patterns. Thesis, Massachusetts Institute of Technology, 2008.

[14] Aileen P. Wright, Adam T. Wright, Allison B. McCoy, and Dean F. Sittig. The use of sequential pattern mining to predict next prescribed medications. Journal of Biomedical Informatics, 53:73–80, February 2015. doi: 10.1016/j.jbi.2014.09.003.

[15] Jonathan H Chen, Mary K Goldstein, Steven M Asch, Lester Mackey, and Russ B Altman. Predicting inpatient clinical order patterns with probabilistic topic models vs conventional order sets. Journal of the American Medical Informatics Association, 24(3):472–80, September 2016. doi: 10.1093/jamia/ocw136.

[16] Jingcheng Du, Peilin Jia, Yulin Dai, Cui Tao, Zhongming Zhao, and Degui Zhi. Gene2vec: Distributed representation of genes based on co-expression. BMC genomics, 20(Suppl 1):82, February 2019. doi: 10.1186/s12864-018-5370-x.

[17] Emeric Dynomant, Romain Lelong, Badisse Dahamna, Clément Massonnaud, Gaétan Kerdelhué, Julien Grosjean, Stéphane Canu, and Stefan J. Darmoni. Word Embedding for the French Natural Language in Health Care: Comparative Study. JMIR medical informatics, 7(3):e12310, July 2019. doi: 10.2196/12310.

[18] Chin Lin, Yu-Sheng Lou, Dung-Jang Tsai, Chia-Cheng Lee, Chia-Jung Hsu, Ding-Chung Wu, Mei-Chuen Wang, and Wen-Hui Fang. Projection Word Embedding Model With Hybrid Sampling Training for Classifying ICD-10-CM Codes: Longitudinal Observational Study. JMIR medical informatics, 7(3):e14499, July 2019. doi: 10.2196/14499.

[19] Clayton A. Turner, Alexander D. Jacobs, Cassios K. Marques, James C. Oates, Diane L. Kamen, Paul E. Anderson, and Jihad S. Obeid. Word2Vec inversion and traditional text classifiers for phenotyping lupus. BMC medical informatics and decision making, 17(1):126, August 2017. doi: 10.1186/s12911-017-0518-1.

[20] Yongjun Zhu, Erjia Yan, and Fei Wang. Semantic relatedness and similarity of biomedical terms: Examining the effects of recency, size, and section of biomedical publications on the performance of word2vec. BMC medical informatics and decision making, 17(1):95, July 2017. doi: 10.1186/s12911-017-0498-1.

[21] Sabrina Jaeger, Simone Fulle, and Samo Turk. Mol2vec: Unsupervised Machine Learning Approach with Chemical Intuition. Journal of Chemical Information and Modeling, 58(1):27–35, January 2018. doi: 10.1021/acs.jcim.7b00616.

[22] Fatima Zohra Smaili, Xin Gao, and Robert Hoehndorf. OPA2Vec: Combining formal and informal content of biomedical ontologies to improve similarity-based prediction. Bioinformatics, 35(12):2133–2140, June 2019. doi: 10.1093/bioinformatics/bty933.

[23] Tomas Mikolov, Ilya Sutskever, Kai Chen, Greg S Corrado, and Jeff Dean. Distributed Representations of Words and Phrases and their Compositionality. In C. J. C. Burges, L. Bottou, M. Welling, Z. Ghahramani, and K. Q. Weinberger, editors, Advances in Neural Information Processing Systems 26, pages 3111–3119. Curran Associates, Inc., 2013.

[24] Andrew L. Beam, Benjamin Kompa, Allen Schmaltz, Inbar Fried, Griffin Weber, Nathan P. Palmer, Xu Shi, Tianxi Cai, and Isaac S. Kohane. Clinical Concept Embeddings Learned from Massive Sources of Multimodal Medical Data. 1804.01486 [cs, stat], April 2018.

[25] Edward Choi, Mohammad Taha Bahadori, Elizabeth Searles, Catherine Coffey, and Jimeng Sun. Multilayer Representation Learning for Medical Concepts. 1602.05568 [cs], February 2016.

[26] Leland McInnes, John Healy, and James Melville. UMAP: Uniform Manifold Approximation and Projection for Dimension Reduction. 1802.03426 [cs, stat], February 2018.

[27] Radim Řehůřek and Petr Sojka. Software Framework for Topic Modelling with Large Corpora. In Proceedings of the LREC 2010 Workshop on New Challenges for NLP Frameworks, pages 45–50, Valletta, Malta, May 2010. ELRA.

[28] Fabian Pedregosa, Gael Varoquaux, Alexandre Gramfort, Vincent Michel, Bertrand Thirion, Olivier Grisel, Mathieu Blondel, Peter Prettenhofer, Ron Weiss, Vincent Dubourg, Jake Vanderplas, Alexandre Passos, and David Cournapeau. Scikit-learn: Machine Learning in Python. Journal of Machine Learning Research, 12:2825–2830, 2011.

[29] Martín Abadi, Ashish Agarwal, Paul Barham, Eugene Brevdo, Zhifeng Chen, Craig Citro, Greg S. Corrado, Andy Davis, Jeffrey Dean, Matthieu Devin, Sanjay Ghemawat, Ian Goodfellow, Andrew Harp, Geoffrey Irving, Michael Isard, Yangqing Jia, Rafal Jozefowicz, Lukasz Kaiser, Manjunath Kudlur, Josh Levenberg, Dan Mané, Rajat Monga, Sherry Moore, Derek Murray, Chris Olah, Mike Schuster, Jonathon Shlens, Benoit Steiner, Ilya Sutskever, Kunal Talwar, Paul Tucker, Vincent Vanhoucke, Vijay Vasudevan, Fernanda Viégas, Oriol Vinyals, Pete Warden, Martin Wattenberg, Martin Wicke, Yuan Yu, and Xiaoqiang Zheng. TensorFlow: Large-scale machine learning on heterogeneous systems. 2015. Software available from tensorflow.org.

[30] Alistair E. W. Johnson, Tom J. Pollard, Lu Shen, Li-wei H. Lehman, Mengling Feng, Mohammad Ghassemi, Benjamin Moody, Peter Szolovits, Leo Anthony Celi, and Roger G. Mark. MIMIC-III, a freely accessible critical care database. Scientific Data, 3:160035, May 2016. doi: 10.1038/sdata.2016.35.

[31] Marzyeh Ghassemi, Tristan Naumann, Peter Schulam, Andrew L Beam, Irene Y Chen, and Rajesh Ranganath. Practical guidance on artificial intelligence for health-care data. The Lancet Digital Health, 1(4):e157–e159, August 2019. doi: 10.1016/S2589-7500(19)30084-6.

